# Preventable Deaths involving Sepsis in England and Wales, 2013-2022: A Systematic Case Series of Coroners’ Reports

**DOI:** 10.1101/2023.08.11.23293989

**Authors:** Jessy Jindal, David Launer, Harrison S France, Molly Hey, Kaiyang Song, Clara Portwood, Georgia Richards, Francesco Dernie

## Abstract

**Purpose:** Coroners’ Prevention of Future Death (PFDs) reports are an under-utilised resource to learn about preventable deaths in England and Wales. We aimed to identify sepsis-related PFDs and explore the causes and concerns in this subset of preventable sepsis deaths.

**Methods:** 4305 reports were acquired from the Courts and Tribunals Judiciary website between July 2013 and November 2022, which were screened for sepsis. Demographic information, coroners concerns and responses to these reports were extracted and analysed, including a detailed paediatric subgroup analysis.

**Results:** 265 reports (6% of total PFDs) involved sepsis-related deaths. The most common cause of death in these reports was “sepsis without septic shock” (42%) and the most common site of infection was the respiratory system (18%) followed by gastrointestinal (16%) and skin (13%) infections. Specific pathogens were named in few reports (27%). Many deaths involved multimorbid patients (49%) or those with recent surgery (26%). Coroners named 773 individual concerns, the most frequent were: a failure to keep accurate records or notes (28%), failure in communication or handover (27%) or failure to recognise risk factors or comorbidities (20%). Paediatric cases frequently reported issues with sepsis screening tools (26%). Sepsis PFDs resulted in 421 individual reports being sent, of which 45% received no response. Most organisations who did respond acknowledged concerns and initiated a new change (74%).

**Conclusion:** Sepsis-related PFDs provide valuable insights into preventable causes of sepsis and identify important sources of improvement in sepsis care. Wider dissemination of findings is vital to learn from these reports.

## Introduction

Sepsis is defined by the World Health Organisation (WHO) as the “life-threatening organ dysfunction caused by a dysregulated host response to infection”, in line with international consensus definitions [1, 2]. Data from the Global Burden of Disease survey estimated that in 2017 there were 48.9 million cases and 11 million sepsis-related deaths worldwide [3]. Estimates of the numbers of deaths related to sepsis are difficult to make and interpret as they rely on the accurate recognition and coding of sepsis among patients. The UK Sepsis Trust estimates that in 2017-2018 there were around 200,000 cases of sepsis in the UK, with around 52,000 associated deaths [4].

The worldwide burden of preventable morbidity and mortality from sepsis is significant [4]. One analysis of 568 patients admitted to hospital with established sepsis in the USA found that 3.7% of sepsis-associated deaths were definitely or moderately preventable when reviewed by clinicians [5]. Despite recognition of the importance of early identification and prompt treatment of sepsis, little is known about the number of preventable deaths related to sepsis, and what factors, systems, or aspects of healthcare fail in practice leading to such deaths.

Since 1984, coroners in England and Wales have had a duty to report and communicate a death where the coroner believes that action should be taken to prevent future deaths [6]. These reports, named Prevention of Future Deaths (PFDs), are mandated under Paragraph 7 of Schedule 5, Coroners and Justice Act 2009, and regulations 28 and 29 of The Coroners (Investigations) Regulations 2013 [7, 8]. These reports are sent to addressees who must then respond within 56 days, and are now used as an official source of data for the NHS Patient Safety Strategy [9]. However, there is no mechanism for systematically synthesizing PFDs to identify and disseminate trends and learnings from coroners’ concerns and actions taken or proposed in their responses.

Studies have started to create case series of PFDs that can be analysed to identify common themes and lessons to be learned, but have yet to examine sepsis [10]. A detailed analysis of PFDs implicating sepsis could highlight important lessons for clinical practice and policy making to prevent avoidable sepsis-related deaths. The aim of our study was to conduct a systematic case series of PFDs to identify sepsis-related deaths, synthesise the concerns raised by coroners, and explore the responses of individuals or organisations to whom these PFDs were addressed.

## Methods

### Study Design

A systematic case series of Prevent Future Death (PFD) reports was designed and the study protocol was preregistered on an open repository [11].

### Data collection, screening, and eligibility

Data were acquired from the Courts and Tribunals Judiciary website [12] on 16^th^ November 2022 using a computer programme called a web scraper designed by a study author (FD) and made openly available [13], similar to the Preventable Deaths Tracker (https://preventabledeathstracker.net/). The web scraper automatically downloaded all published PFD portable document formats (pdfs) from the Judiciary website and searched for pre-determined keywords. The keywords chosen were ‘sepsis’, ‘infection’, ‘septic’, ‘shock’, ‘infected’, ‘infective’ and the positive control word ‘coroner’ (to identify documents which couldn’t be automatically screened for the keywords). The code identified and downloaded 4305 pdfs, of which 4205 were PFDs and the remainder were mis-labelled response letters. Nearly half (47.8%; n=2010) of the pdfs were readable by code and eligible for primary inclusion and were screened by at least one author (JJ, FD) for validation purposes (Supplementary Figure S1). The remaining 2295 pdfs were independently screened by at least two study authors (JJ, DL, HF, MH, KS, CP). Cases were included if sepsis caused or contributed to death. Sepsis was defined as per the WHO definition: “life threatening organ dysfunction caused by a dysregulated host response to infection”. Included cases either had documented sepsis explicitly as a cause of death or had very strong evidence of sepsis (i.e., multi-organ failure in the context of a severe infection). Cases where death was caused by other severities of infection were excluded. Any ambiguities regarding inclusion of cases were discussed with and resolved by the senior investigator (FD).

### Data extraction

Two authors (JJ, DL) manually extracted case demographics, causes of death and morbidity, named microorganism(s) causing infection, antibiotic(s) given if named, cause of infection (origin), issues with care for comparison with the Royal College of Emergency Medicine (RCEM) audit [14], sepsis risk factors, coroners’ concerns and response from recipients receiving PFDs. Data regarding the geographic distribution of all PFDs was extracted from the Preventable Deaths Tracker [15]. We extracted the numbers of sepsis-related deaths from the Office for National Statistics (ONS) [16], to determine the number of sepsis deaths written up into PFDs per year.

### Analysis

The number of included sepsis-related PFDs, their rates as a proportion of all PFDs, and ONS mortality data were calculated over time. Medians and interquartile ranges (IQRs) were calculated for continuous variables (e.g., age) and frequencies were reported for categorical variables (e.g., sex, location of death, coroner area of jurisdiction).

We calculated the years of life lost (YLL) [17] for each case (where age was reported) by extracting their remaining life expectancy from the Office for National Statistics (ONS) cohort life tables [18]. Two investigators (JJ, DL) assigned the International Statistical Classification of Disease and Related Health Problems 11^th^ Revision (ICD-11) [19] numeric codes for the causes of death to each PFD. To examine geographical variation, we graphed both the absolute counts and rates of sepsis-related PFDs per all PFDs in each coroner area mapped to the standard regions of England and Wales. To evaluate reporting of issues with initial sepsis-related care reported in PFDs, information was extracted from the Royal College of Emergency Medicine (RCEM) sepsis audit and reported as a percentage of total cases.

To collate and evaluate concerns raised by coroners, we classified and identified repeated themes using directed content analysis [20]. This allowed us to highlight cases involving previously recognised concerns, and to explore concepts, drawing similarities and disparities from the data. A paediatric (age <18 years) subgroup analysis was undertaken to explore whether particular concerns regarding this age group were more prevalent.

To calculate response rates to PFDs, we used the 56-day legal requirement [7], to classify responses as ‘on time’, ‘late’, or ‘unspecified’ (where a date was not present). We calculated the average response rate and frequency for individuals and organisations. Responses from organisations in receipt of sepsis-related PFDs were synthesised and classified by type of change initiated using content analysis.

### Software

We used Datawrapper [21] to produce bar charts, pie charts and the choropleth map. We used R (version 4.1.1), including the pdftools package, to create the web scraper that downloads and screens PFDs, which is openly available [13].

### Ethics

We are using publicly available information, for which ethics committee approval is not required.

### Funding

No funding has been obtained to undertake this study.

## Results

### Demographics

There were 265 sepsis-related PFDs, related to 265 deaths between July 2013 and 16 November 2022 in England and Wales (6.3% of all PFDs). The rate of sepsis-related PFDs remained roughly constant from 2013 (8.2%) to 2022 (6.4%) (Supplementary Table S1). Using ONS data for deaths where “sepsis was the underlying cause of death”, a median of 0.13% (IQR: 0.11-0.15%) of all sepsis-related deaths in England and Wales were written into PFDs each year (Supplementary Table S1).

The median age at death was 69 years old (IQR: 38 to 80 years; n=148), with a median of 17.5 years of life lost per person (IQR: 9.8 to 47.1; n=148). 31 cases (11.7%) were aged less than 18 years old at the time of death. Just over half (54.7%; n=265) of those who died were male (Figure 1).

**Fig 1.**
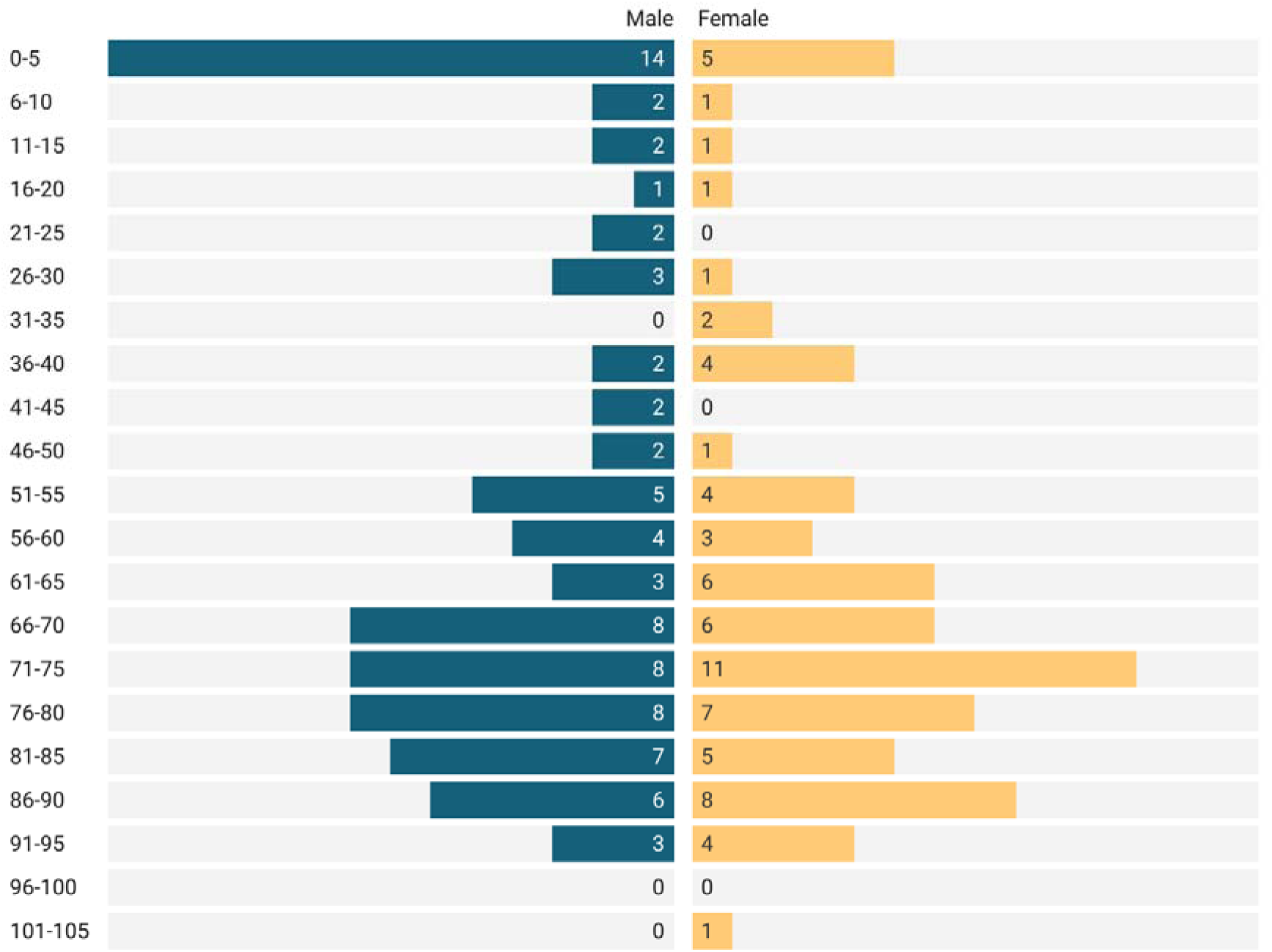
Age distribution of included fall-related PFD cases, by sex.

There were 265 sepsis PFDs across 59 Coroner’s Areas (Supplementary Table S2). The largest number were from North-West England (25.7%) followed by London (18.5%) and Yorkshire and the Humber (11.3%) (Supplementary Table S3, Figure 2a). The highest number of sepsis-related PFDs as a proportion of total PFDs from each region came from Yorkshire and the Humber (10.1%), followed by North-West England (8.0%) and London (7.6%) (Supplementary Table S3, Figure 2b).

**Fig 2.**
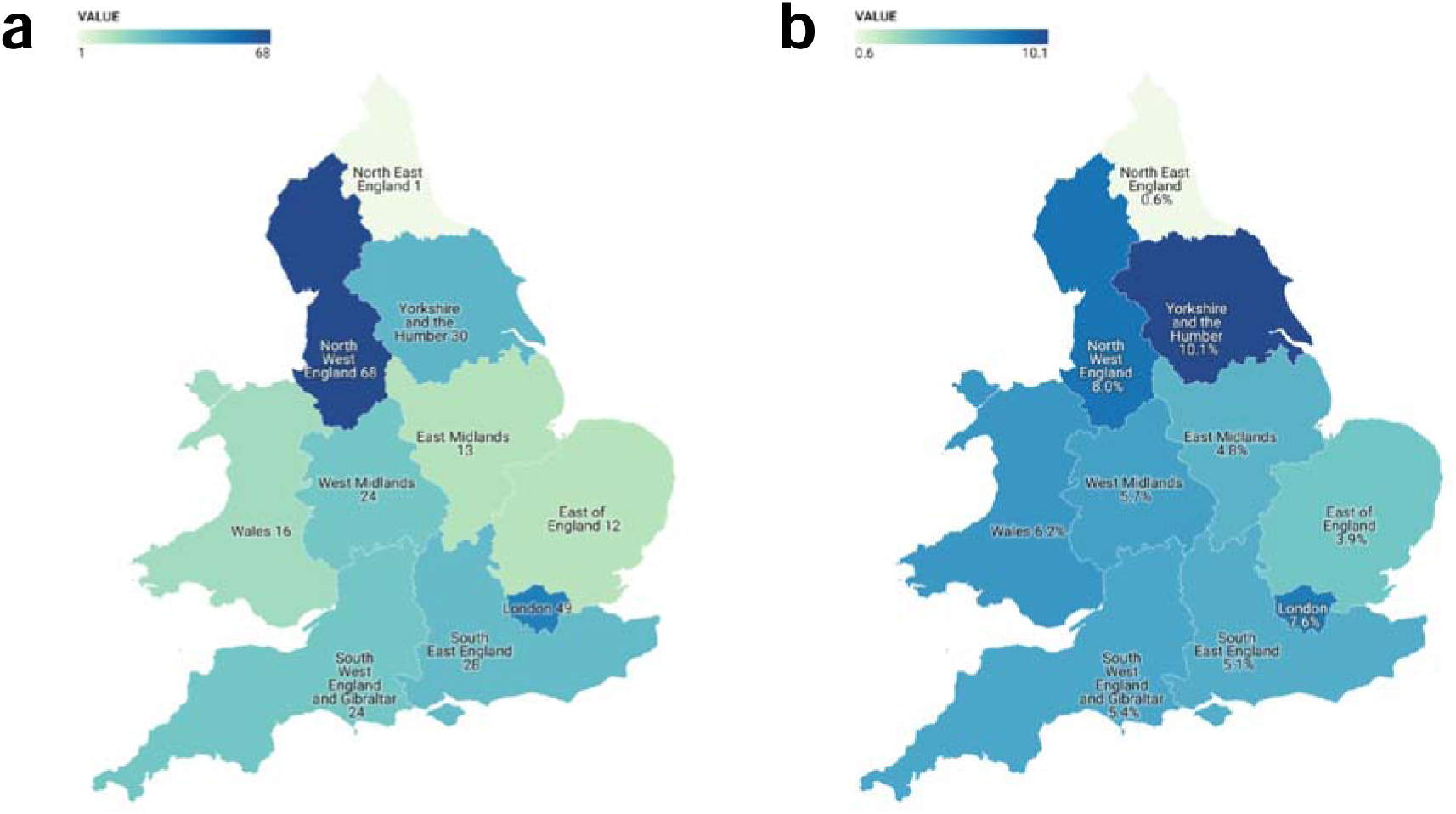
a) Raw number of sepsis-related PFDs published per region, 2013-2022. b) Number of sepsis-related PFDs published per region expressed as a percentage of all PFDs published by that region, 2013-2022.

### Causes of death and sepsis risk factors

On the Judiciary website, the majority (75.1%) of sepsis-related PFDs were classified under the ‘Hospital Death’ report type (Supplementary Table S4). Other common categories of classification were ‘Community Healthcare and Emergency Services’ (11.7%), ‘Care Home related Deaths’ (9.4%), and ‘Child’s death’ (6.8%).

After categorisation using ICD-11 codes, the two most common causes of death were ‘Sepsis without septic shock’ (41.7%) and multi-organ failure (21.3%). Other common causes were ‘sepsis with septic shock’, pneumonia, and urinary tract infections (Supplementary Table S5).

The most frequent site of infection was the respiratory system (18.5% cases), followed by gastrointestinal (GI) infections (15.9%) and skin infections (12.8%) of which a large portion (20/34) were pressure sores (Table 1).

**Table 1.**
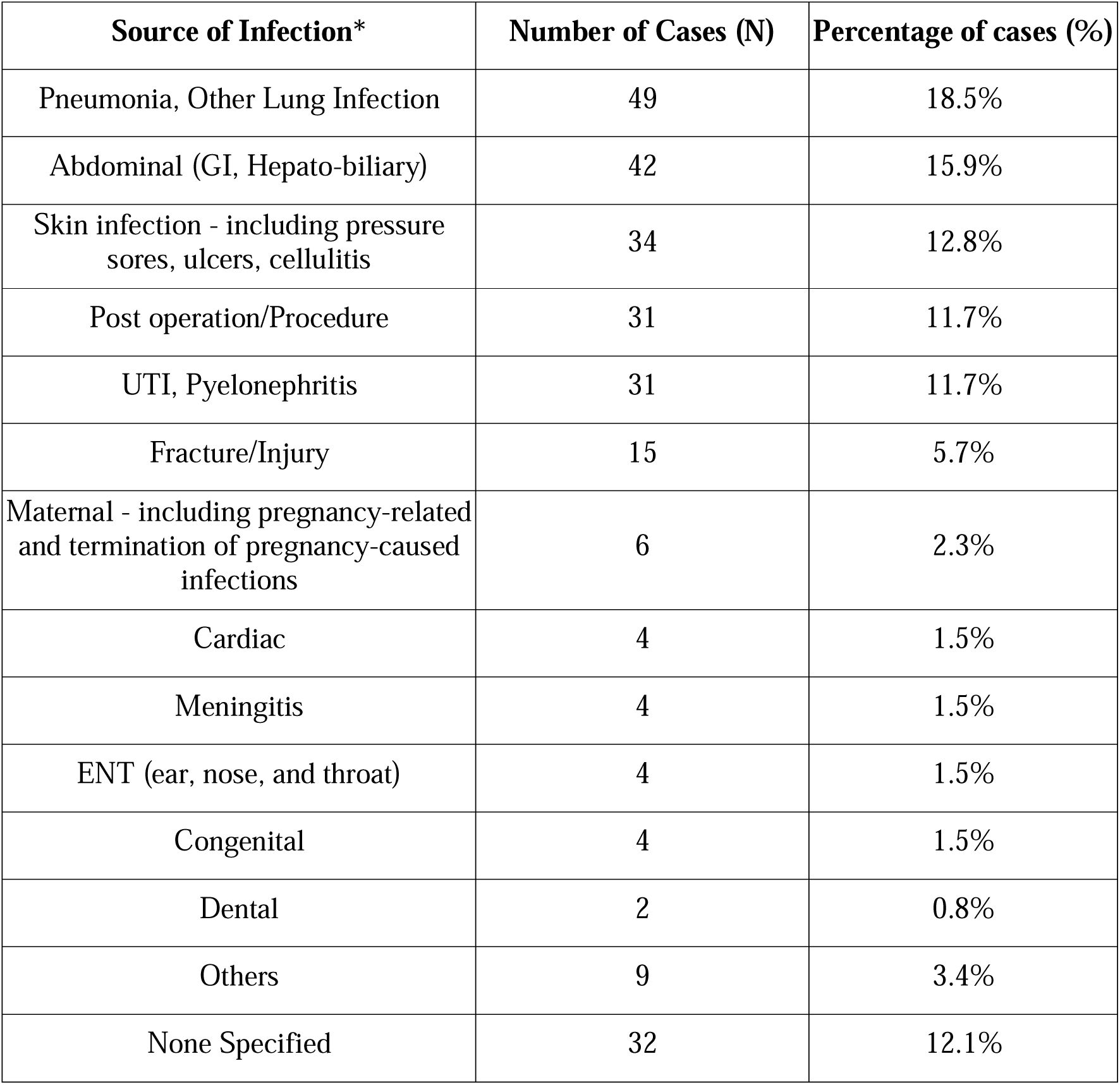
Source of infection as outlined in PFD report. *Note: Some cases had more than one site of infection specified.

Specific pathogens were named in 27.2% of PFDs (Supplementary Table S6). Of these 90.2% were bacteria, the most common being E. coli (22.2%), Group A Streptococcus (13.9%), and N. meningitidis (8.3%). Viral (5.6%) and fungal (4.2%) pathogens were specifically mentioned in relatively few PFDs.

Of the risk factors for sepsis extracted from PFDs, multimorbidity (49.4%) and recent surgery (26.0%) were most prevalent. Immunosuppression from drug therapy, neutropenia, Human immunodeficiency virus (HIV) positivity and genetic immunodeficiency were all present in <5% cases (Supplementary Table S7).

### Coroner’s Concerns

Coroners expressed 773 individual concerns in the 265 sepsis-related PFDs. Using content analysis, these concerns were categorised into 18 themes. Concerns were most often related to a failure to keep accurate records or notes (27.6%), failure in communication or handover (27.2%), and a failure to recognise risk factors or comorbidities (19.6%). Of note, 206 PFDs mentioned a concern relevant to a diagnosis other than sepsis (77.7%), and this was the sole concern in 60 (22.6%) of the included PFDs (Supplementary figure S8). Examples of concerns from the most common themes are included in Table 2.

**Table 2.** Selected examples of specific concerns raised by coroners from the three most common concern themes. *Prior to case numbers being introduced, only dates are available.

| Theme | Case number | Example Concerns |
| --- | --- | --- |
| **Failure to keep accurate records or notes (n=73, 27.6%)** | 2020-0110 | “The quality of the documentation was not always of a good standard and part of the reason why his catheter was incorrectly believed to be a long-term catheter and left in, [ultimately leading to urosepsis]” |
|  | 15/12/2015* | “Mrs (redacted) should have undergone hourly observations. This did not happen. There was reference to observations being undertaken at 00.30 but these were not recorded” |
| **Failure in communication or handover (n=72, 27.2%)** | 2018-0329 | “nurse [...] bumping into (redacted) believing he did not look well enough to go to the toilet. He was not aware of (redacted) as no handover had been given. [...] Failure of clinical staff to handover clinical concerns to each other and to medical staff including face to face handovers” |
|  | 2019-0446 | “When the plan [for a procedure] changed, there was no communication between the interventional radiologist and the hepatologists. Even if it had not changed the |
|  |  | plan, Mr (redacted)’s management would have benefited from a robust discussion between the specialists in these two fields, and an accurate record of the decision making” |
| **Failure to recognise risk factors or comorbidities (n=52, 19.6%)** | 2022-0129 | “Mr (redacted) was frail and vulnerable with very poor swallow. When prescribing the clinicians did not recognise or have a system to flag up the need for liquid antibiotics rather than tablet antibiotics. This led to him not being able to commence antibiotics on the day he was identified as needing them” |

The cases were compared to the issues identified in the RCEM audit on severe sepsis deaths; the most frequent problems leading to preventable deaths were lack of senior review (19.3%, n=51), issues with education and training (17.4%, n=46), issues with administration of antibiotics (14.3%, n=38), and issues with NEWS scores/sepsis screening (12.1%, n=32) (Supplementary figure S9).

Coroners sent 421 PFDs to organisations (Supplementary Table S10). NHS trusts or hospitals received the most reports (37.5%), and 42% of their responses were overdue (i.e., no response within 56 days of receiving the report). The response rate varied widely between organisations: 37% responded early or on time, 14% responded late, and 45% remained overdue as of the time of writing. 4% of reports were responded to but not dated.

When PFDs were responded to, responses from organisations were categorised thematically, into whether they acknowledged the coroner’s concern and initiated a new change (74% overall), acknowledged the concern but argued that a pre-existing solution was adequate (20%), or responded but did not acknowledge or agree with the concern (6%) (Figure 3a, Supplementary Table S11). Of these, the most common new changes were introducing a new protocol, pathway, or guidance document (55%); starting an audit or investigation (37%); or improving training (37%) (Figure 3b, Supplementary figure S12).

**Fig 3.**
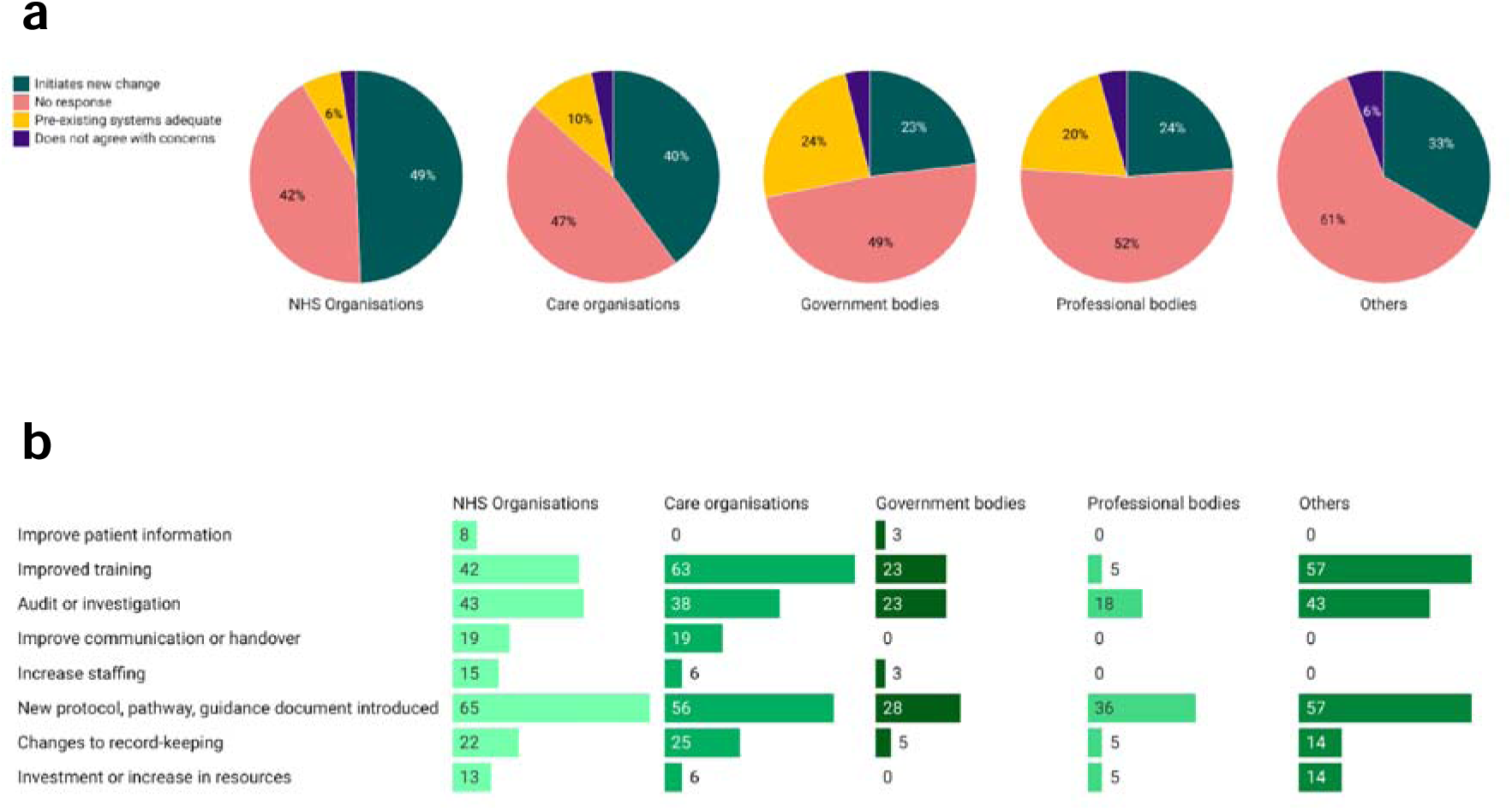
a) Initial higher-order response classification by organisation type. b) Classification of changes initiated by organisations (as a percentage of all responses) in response to coroner concerns.

### Paediatric deaths related to sepsis

31 PFDs related to paediatric deaths (<18 years old). Of these, 23 had specific ages mentioned, of which the median age was 0.9 years (11 months of age), and mean age was 3.0 years. The median years of life lost for this paediatric group was 87 years per person (IQR 82.7-87.0, n=23). The majority of paediatric cases were male (74%).

Sepsis risk factors in the paediatrics group were not frequently reported. The most common causes of death were ‘sepsis without septic shock (45%), ‘sepsis with shock’ (16%), meningococcal disease (10%), and bacteraemia (10%) (Supplementary Figure S13). Pathogens were named in 21/31 (68%) of paediatric PFDs. The most common was E.coli (29%) (Supplementary Table S14). Respiratory infections were the most frequent infection source and renal/urinary tract infections were also common (Supplementary Table S15). There was a high rate of disseminated septic shock without a clear source of infection (35.5% of paediatric cases).

When compared to the RCEM audit of severe sepsis deaths, issues identified were similar, the most common being issues with education and training for sepsis recognition, mentioned in 39% of reports. (Supplementary Table S16). The two most common concerns identified by coroners were failure to use sepsis screening tools (26%) and failure to provide appropriate treatment (26%) (Supplementary Table S17). Only 16% of reports mentioned a failure of accurate record keeping, which was the most common identified concern in adult reports (27%).

Coroners sent 53 reports to organisations, the most common recipient being NHS trusts (25% of reports sent) (Supplementary Table S18). The timing of responses was comparable to all PFDs: the response rate was 40%, with 23% being early or on time, 15% late, and 60% overdue (Supplementary Table S19). When reports were responded to, 62% of responses acknowledged coroners’ concerns and initiated a new change, 33% argued that a pre-existing solution was adequate, and 5% did not agree with the concerns raised (Supplementary Table S20). Out of any new changes initiated, new protocols, pathways, or guidance documents were mentioned in 52% of responses to reports (Supplementary Table S21).

## Discussion

### Summary of Results

We identified 265 reports involving sepsis where coroners believed that actions should be taken to prevent similar deaths. On average, 17.5 years of life were lost for every sepsis-related PFD. Where identified, the most common septic sources were the lungs, GI tract, skin infections, procedure-related infections, or the urinary tract. The most common organisms implicated were bacteria, although specific pathogens were named in relatively few reports (27.2%). Coroners raised nearly 800 concerns including failures to keep accurate records, failures in communication, or a failure in recognising a sepsis risk factor. Despite a legal requirement to respond within 56 days, 45% of organisations had not responded by the time of writing, or their responses have not been published. Where they have responded, the large majority have introduced a new change to prevent similar sepsis deaths from reoccurring.

31 PFDs related to paediatric deaths (<18 years old), of which the median age was 0.92 years (11 months of age), and mean age was 3 years. Disseminated sepsis without a clear source of infection was especially common in paediatric cases, although respiratory tract infections remained the most frequent source. Pathogens were named in a high proportion of paediatric cases (68%) compared to total group (27.2%). Accurate record keeping was an underrepresented concern, while education and training was cited frequently.

### Comparisons with existing literature

This is the first study to focus on sepsis-related PFDs, or PFDs related to any infectious cause of death. Previous studies of PFDs investigating deaths relating to medicines, opioids, and falls have also identified a poor response rate and lack of standardisation in PFD content [13, 22, 23]. These concerns were also identified in a House of Commons Justice Select Committee [24].

Comparing our data to those from studies covering all sepsis deaths can show factors overrepresented in PFD reports. In UK Sepsis Trust data, pneumonias (50%), urinary tract infections (20%), abdominal infections (15%) and skin, soft tissue, bone and joint infections (10%) were identified as the largest sources of infection for sepsis cases [4]. Whilst these causes were seen at a high rate in our sepsis PFDs, we did not find any one source to account for more than 20% of cases. We also found a higher rate of procedure/surgery related causes (11.7%) whilst “device-related infection” (the only iatrogenic source of sepsis included in their data) only formed 1% of cases in the Sepsis Trust data; this perhaps reflects that iatrogenic sources of infection result in more ‘preventable’ cases of sepsis deaths.

The RCEM Audit 2016/17 identified factors in the development of sepsis presenting to 196 emergency departments (EDs). The RCEM data reports statistics on whether standard care packages for sepsis are met in EDs, namely the ‘SEPSIS-6’ elements and whether these were completed within goal times. We compared our cases where possible to their summary of recommendations of care, although coroners did not consistently comment on these aspects of care. Lack of senior review was an issue in 38% of cases reviewed by the RCEM, but only appeared in 19.3% of PFDs. RCEM reports that 96% of EDs provide sepsis education, but we found that 17.4% of coroners’ concerns surrounded problems with education and training. RCEM data showed only 44% of patients receive antibiotics within an hour of arrival, and we found administration of antibiotics contributed to preventable deaths in 14.3% of our cases.

Common risk factors identified among our patients included multimorbidity and recent surgery. Increased risk of death from sepsis and organ failure amongst multimorbid patients is well characterized in the literature [25]. The Sepsis Trust reports around 1% risk of sepsis in post-operative patients in the UK, while post-operative risk of sepsis in other countries’ literature ranges from 1.3% in Australia [26] to 3.9% in the USA [27] to 4.4% in Taiwan [28]. Current early detection systems appear to do a poor job at predicting postoperative sepsis - a study looking at sepsis in post-operative colorectal patients found that the recommended Quick Sequential Organ Failure Assessment (Q-SOFA) score was “little better than a coin toss” at identifying signs of sepsis in the first 48 hours [29].

### Strengths and Limitations

This study builds on prior research looking at subgroups of PFDs [22, 30–34], and addresses concerns specific to sepsis. Using reproducible and prespecified data collection methods, we extracted and analysed information from 265 sepsis-related PFDs. These are a small fraction of sepsis-related deaths in the UK but are an important source of information for preventable deaths as addressed in the NHS Patient Safety Strategy [9].

There are important limitations in using information from PFDs. There is established large inter-regional and inter-coroner variation in the writing of PFDs [35]. The PFDs that are written are not subject to any routine quality assurance or auditing, so that the reports depend on the working practices of the individual coroner - both in terms of content included, and in subjectivity of the concerns identified. PFDs do not consistently include demographic data such as age, ethnicity, or location of death. Another limitation was the relative subjectivity in classifying reports as “sepsis-related deaths”, however we endeavoured to address this by only including reports where sepsis was explicitly mentioned, or where there was a reasonable description of multi-organ failure related to sepsis. This is compounded by the variable use of the term in general. Other limitations relate to the responses by organisations - we could only address those uploaded to the judiciary website while there may be more that had been written, and we could not verify that the changes proposed in these responses were implemented in practice.

### Implications for policy and practice

An estimated 245,000 patients are admitted to hospitals in the UK every year with sepsis [3], with mortality estimated by the UK Sepsis Trust to be 28.9% in England [4]. Risk of death remains considerable after leaving hospital, with 15% of patients who survive sepsis in hospital dying within the next year [36]. Sepsis remains a life-threatening condition with optimal care, but the coroners’ reports explored within this study show how numerous factors related to the care of a patient with sepsis can, when they go wrong, lead to a preventable death.

The influence of human factors [37] on patient safety is increasingly being recognised in the NHS, including in the NHS Patient Safety Strategy [9]. These include issues with communication or handover, which coroners raised in more than a quarter (27.2%) of sepsis PFDs. However, only 13% of responses to PFDs initiated changes in this area, suggesting there is still a disconnect between the identification of human factors-related issues and enacting change to prevent similar issues occurring again. PFD findings help identify common system or organisational issues which lead to human errors in sepsis management, and their wider dissemination could help drive quality improvement in this area nationally. These findings could supplement existing sepsis education programmes which are effective in improving knowledge around sepsis and patient outcomes [38].

Analysis of sepsis-related PFDs highlighted a number of common clinical contexts where coroners raised concerns. One such scenario was recent surgery, which was present in 26% of PFDs. The issue of surgical site infections (SSIs), and the risk of developing subsequent sepsis, is a significant cause of morbidity and mortality globally [39]. A 2019 guideline by the National Institute for Health and Care Excellence (NICE) [40], outlines pre-operative (hygienic preparation and in some circumstance prophylactic antibiotics), intra-operative (sterility and appropriate surgical techniques) and post-operative (wound management and identification of infection) steps to prevent SSIs. The PFDs in our report raise concerns with various aspects of post-operative care including adequate discharge planning and regular wound care. Cohort studies have shown that post-operative sepsis is associated with increased post-discharge mortality [26], suggesting that in addition to reinforcing SSI prevention training, healthcare professionals should be alert to this vulnerable population in the community.

Almost half (49.4%) of cases were explicitly described as multi-morbid or were clearly multimorbid based on the international definition of the presence of two or more long-term medical conditions [41]. Multimorbidity is known to lead to worse outcomes from sepsis [25]. Given the risk of quicker deterioration, and poorer response to therapy, these patients should be identified early by clinicians and paid close attention to prevent deaths from sepsis. Less common sites of infection causing eventual sepsis were also highlighted by PFDs, including pressure sores or ulcers - identified in 7.6% of our PFDs (n=20), but not mentioned anywhere in the Sepsis Trust’s Surviving Sepsis Manual [4]. Pressure ulcers affect 700,000 patients in the UK each year [42] and have a sepsis-associated mortality rate as high as 40% [43], indicating a vulnerable group of patients that may need special attention when looking for signs of infection.

Paediatric deaths identified certain concerns not seen in adult cases – in particular, multiple reports cited deficiencies in paediatric sepsis screening tools. A recurring concern involved standardisation of Paediatric Early Warning Scores across hospitals – a lack of consistency could contribute to low compliance, especially amongst locum or part-time staff. Other issues raised included a lack of education in recognition of uncommon paediatric presentations like pancreatitis.

## Conclusions

Sepsis-related PFDs provide valuable insights into the processes and errors which lead to preventable sepsis deaths. Wider dissemination of their findings can support quality improvement initiatives and education programmes to improve sepsis care and patient safety.

## Statements and declarations

### Author contributions

FD conceptualised the project and wrote the code used to download and screen the PFDs. GCR created the Preventable Death Tracker and established the methodology to analyse PFDs. FD wrote the study protocol. All authors were involved in screening the PFDs for inclusion. DL and JJ performed the data extraction and data synthesis and wrote the manuscript. All authors were involved in interpretation of the results. All authors have read and endorse the findings of the final manuscript.

## Data Availability

All data produced in the present study are available upon reasonable request to the authors

