## Supplementary Material for "Preventable Deaths involving Sepsis in England and Wales, 2013-2022: A Systematic Case Series of Coroners’ Reports"

**Title**

Dr Francesco Dernie

Oxford University Hospitals NHS Foundation Trust, Oxford, OX3 9DU, UK.

**Supplementary Figure S1.** Flow diagram of inclusion and exclusion of cases downloaded from the Judiciary website.

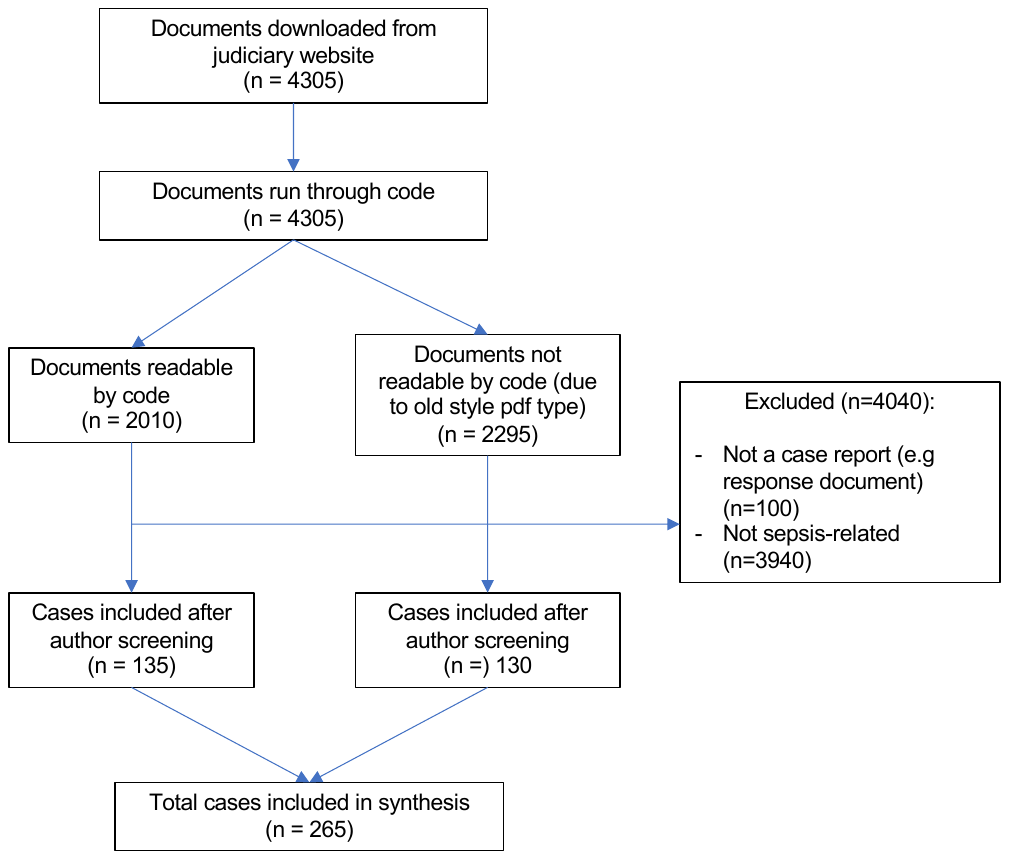

**Supplementary Table S1** Number of sepsis-related PFDs by year, including as a percentage of all PFDs for that year and as a percentage of all sepsis-related deaths for that year as documented by the Office for National Statistics (ONS)

| **Year** | **Total number of PFDs** | **Number of sepsis-related PFDs** | **Rate of sepsis-related PFDs** | **Number of ONS sepsis deaths** | **Percentage of sepsis PFDs vs ONS sepsis deaths** |
| --- | --- | --- | --- | --- | --- |
| 2013 | 135 | 11 | 8.15% | 22,967 | NA* |
| 2014 | 614 | 24 | 3.91% | 22,826 | 0.11% |
| 2015 | 492 | 22 | 4.47% | 24,784 | 0.09% |
| 2016 | 398 | 28 | 7.04% | 24,973 | 0.11% |
| 2017 | 446 | 35 | 7.85% | 23,709 | 0.15% |
| 2018 | 386 | 31 | 8.03% | 23,185 | 0.13% |
| 2019 | 596 | 33 | 5.54% | 21,458 | 0.15% |
| 2020 | 301 | 23 | 7.64% | 19,324 | 0.12% |
| 2021 | 476 | 35 | 7.35% | 21,947 | 0.16% |
| 2022 | 361 | 23 | 6.37% | NA† | NA† |
| **Total** | 4205 | 265 | 6.30% | 205173 | 0.13% |
| **Median (IQR)** | 422 (367.25-488) | 26 (23 – 32.5) | 7.19% (5.75% - 7.80%) | 22,967 (21,947 - 23,709) | 0.13% (0.11 – 0.15%) |
| * Data only from July 2013 onwards - percentages not representative for 2013 | | | | | |
| † ONS figures for Sepsis deaths only up to 2021, used “Sepsis was mentioned anywhere on the death certificate” | | | | | |

**Supplementary Table S2.** Classification of sepsis-related Prevention of Future Death reports (PFDs) according to coroner area described in the report.

| **Coroner Area** | **Number of PFDs (n=265)** |
| --- | --- |
| Avon | 5 |
| Bedfordshire & Luton | 4 |
| Berkshire | 1 |
| Birmingham and Solihull | 6 |
| Black Country | 6 |
| Blackpool & Fylde | 3 |
| Brigend, Glamorgan Valleys and Powys | 1 |
| Buckinghamshire | 1 |
| Camarthenshire and Pembrokeshire | 1 |
| Cardiff & the Vale of Glamorgan | 1 |
| Cornwall and the Isles of Scilly | 5 |
| Derby and Derbyshire | 1 |
| Dorset | 2 |
| Gateshead & South Tyneside | 1 |
| Gloucestershire | 6 |
| Greater Manchester South | 7 |
| Isle of Wight | 1 |
| Kent (Central & South East) | 1 |
| Lancashire and Blackburn with Darwen | 2 |
| Leicester City & South Leicestershire | 4 |
| Liverpool and Wirral | 3 |
| London (City) | 1 |
| London (East) | 11 |
| London (North) | 1 |
| London (South) | 3 |
| London (West) | 3 |
| London Inner (North) | 18 |
| London Inner (South) | 12 |
| Manchester (City) | 6 |
| Manchester (North) | 5 |
| Manchester (South) | 32 |
| Manchester (West) | 10 |
| Mid Kent and Medway | 4 |
| Milton Keynes | 2 |
| Norfolk | 5 |
| Northampton | 2 |
| North East Kent | 2 |
| North Wales (East and Central) | 5 |
| North West Kent | 4 |
| Nottinghamshire | 8 |
| Plymouth Torbay and South Devon | 5 |
| Portsmouth & South East Hampshire | 1 |
| Powys, Bridgend & Glamorgan Valleys | 1 |
| Shropshire, Telford & Wrekin | 1 |
| South Lincolnshire | 1 |
| South Wales Central | 7 |
| South Yorkshire (East) | 3 |
| South Yorkshire (West) | 3 |
| Staffordshire South | 1 |
| Stoke-on-Trent & North Staffordshire | 8 |
| Suffolk | 3 |
| Sunderland | 2 |
| Surrey | 12 |
| Swansea, Neath and Port Talbot | 2 |
| West Sussex | 4 |
| West Yorkshire (East) | 9 |
| West Yorkshire (West) | 3 |
| Wiltshire & Swindon | 1 |
| Worcestershire | 2 |

**Supplementary Table S3.** Classification of sepsis-related Prevention of Future Death reports (PFDs) according to administrative regions of England and Wales.

| **Region** | **Number of sepsis-related PFDs (n=265)** | **Distribution of PFDs across different regions** | **PFDs per region (n=4205)** | **Rate of sepsis PFDs by region** |
| --- | --- | --- | --- | --- |
| East Midlands | 13 | 4.9% | 272 | 4.8% |
| East of England | 12 | 4.5% | 310 | 3.9% |
| London | 49 | 18.5% | 649 | 7.6% |
| North East England | 1 | 0.4% | 156 | 0.6% |
| North West England | 68 | 25.7% | 848 | 8.0% |
| South East England | 28 | 10.6% | 547 | 5.1% |
| South West England and Gibraltar | 24 | 9.1% | 446 | 5.4% |
| Wales | 16 | 6.0% | 259 | 6.2% |
| West Midlands | 24 | 9.1% | 422 | 5.7% |
| Yorkshire and the Humber | 30 | 11.3% | 296 | 10.1% |

**Supplementary Table S4.** Number of cases classified under each Judiciary website category. Note that individual PFDs may be tagged with multiple categories.

| **Category** | **Number of sepsis related PFDs** | **Percentage of total sepsis-related PFDs** |
| --- | --- | --- |
| Hospital Death | 199 | 75.09% |
| Community Healthcare +/- Emergency Services | 31 | 11.70% |
| Care Home Related Death | 25 | 9.43% |
| Child’s Death | 18 | 6.79% |
| Other Related Death | 14 | 5.28% |
| Wales (2019 onwards) | 7 | 2.64% |
| Emergency services related deaths (2019 onwards) | 6 | 2.26% |
| Alcohol, Drugs | 5 | 1.89% |
| Mental Health | 4 | 1.51% |
| Accident at Work and Health and Safety related deaths | 2 | 0.75% |
| State Custody Related Deaths | 1 | 0.38% |

**Supplementary Table S5.** ICD-11 codes for cause of death accounting for ≥0.5% cases (Section 1A of death certificate).

| **ICD-11 coding for cause of death** | **Number of cases (N)** | **Percentage of cases (%)** | **Specific** |
| --- | --- | --- | --- |
| 1G40 | 96 | 41.7% | Sepsis without septic shock |
| MG4A | 49 | 21.3% | Multi-Organ failure |
| 1G41 | 15 | 6.5% | Sepsis with Septic Shock |
| CA40, CA40.Z | 13 | 5.7% | Pneumonia |
| GC08.Z | 6 | 2.6% | UTI |
| 1C1C.20, 1C1C, 1C1C.2 | 4 | 1.7% | Meningococcal disease |
| GB60, GB60.Z | 4 | 1.7% | Acute kidney failure |
| 1H0Z | 2 | 0.9% | Infection, unspecified |
| 8B24 | 2 | 0.9% | Hypoxic-ischaemic encephalopathy |
| BA41.Z | 2 | 0.9% | Acute myocardial infarction |
| BD11 | 2 | 0.9% | Left ventricular failure |
| DC50 | 2 | 0.9% | Peritonitis |
| MA15.0 | 2 | 0.9% | Bacteraemia |
| NA07.6 | 2 | 0.9% | Traumatic subdural haemorrhage |

**Supplementary Table S6.** Specific pathogens identified within PFDs as the causative agent in sepsis. A single PFD may mention more than one pathogen.

| **Pathogen** | **N** | **% of PFDs which named a pathogen** |
| --- | --- | --- |
| **Bacterial** | 65 | **90%** |
| Escherichia .coli | 16 | 22% |
| Group A streptococcus | 10 | 14% |
| Neisseria meningitidis | 6 | 8% |
| Staphylococcus aureus | 4 | 6% |
| Streptococcus pneumoniae | 4 | 6% |
| Streptococcus unspecified | 4 | 6% |
| Group B streptococcus | 2 | 3% |
| Klebsiella | 2 | 3% |
| Pseudomonas aeruginosa | 2 | 3% |
| Beta haemolytic streptococcus, group unspecified | 1 | 1% |
| Clostridium difficile | 1 | 1% |
| Capnocytophagia canimorsus | 1 | 1% |
| Coagulase-negative Staphylococcus | 1 | 1% |
| Enterococcus faecium | 2 | 3% |
| Enterobacter cloacae | 1 | 1% |
| Gram negatives unspecified | 1 | 1% |
| Legionella pneumophilia | 1 | 1% |
| Mixed anaerobes | 1 | 1% |
| Morganella | 1 | 1% |
| MRSA | 1 | 1% |
| Mycobacteria chimaera | 1 | 1% |
| Serratia Marcescens | 1 | 1% |
| Stenotrophomonas matophilia | 1 | 1% |
| **Viral** | 4 | 6% |
| Influenza A/B | 3 | 4% |
| COVID-19 | 1 | 1% |
| **Fungal** | 3 | 4% |
| Aspergillus | 2 | 3% |
| Candida sp. | 1 | 1% |
| **Total** | 72 | 100 |

**Supplementary Table S7.** Prevalence of sepsis risk factors highlighted by coroners in sepsis-related PFDs

| **Risk Factor** | **Number of Cases** | **%** |
| --- | --- | --- |
| Immunosuppression from drug therapy | 9 | 3.40% |
| Neutropenia | 6 | 2.26% |
| HIV | 2 | 0.75% |
| Genetic immunodeficiency | 3 | 1.13% |
| Multi-morbidity | 131 | 49.43% |
| Recent surgery | 69 | 26.04% |

**Supplementary Table S8:** Coroners’ concerns grouped following thematic analysis.

| **Concern** | **N** | **% of cases where concern present** |
| --- | --- | --- |
| Failure to keep accurate records / notes | 70 | 26% |
| Failure in communication or handover | 66 | 25% |
| Failure to recognise risk factors/comorbidities | 52 | 20% |
| Failure to provide appropriate Treatment | 49 | 18% |
| Lack of training | 46 | 17% |
| Failure to escalate | 45 | 17% |
| Failure in timely medical assessment of patient | 39 | 15% |
| Understaffing | 35 | 13% |
| Failure to use sepsis screening tools ie. problems with early warning systems | 32 | 12% |
| Failure to take observations | 30 | 11% |
| Failure to follow relevant pathways, protocols, guidelines or risk assessments once sepsis had been recognised | 22 | 8% |
| Lack of resources | 18 | 7% |
| Poor response time of inpatient medical staff | 17 | 6% |
| Poor discharge planning | 16 | 6% |
| Poor response time of emergency services | 13 | 5% |
| Poor response time of care home staff | 11 | 4% |
| Failure to secure imaging in timely manner/at all | 10 | 4% |
| Concern not relevant to sepsis e.g. DNAR in place was ignored | 202 | 76% |

**Supplementary Table S9:** Concerns raised by coroners in line with the Royal College of Emergency Medicine (RCEM) sepsis audit

| **RCEM category** | **Number of cases** | **% of cases** |
| --- | --- | --- |
| Issues with senior review / escalation | 51 | 19.25 |
| Issues with education and training for early recognition and instigation of optimal care | 46 | 17.36 |
| Issues with administration of antibiotics | 38 | 14.34 |
| Issues with NEWS scores and sepsis screening | 32 | 12.08 |
| Issues with sepsis protocol / sepsis lead | 26 | 9.81 |
| Issues with sepsis protocol / sepsis lead | 24 | 9.06 |
| Issues with sepsis care pathways | 17 | 6.42 |
| Issues with monitoring of parameters including lactate and urine output | 12 | 4.53 |
| Issues with providing patients with written information | 12 | 4.53 |
| Issues with administration of oxygen | 3 | 1.13 |

**Supplementary Table S10**: Organisations and individual who received sepsis-related PFDs, and their response rates according to the statutory requirement of 56 days. Similar individual organisations have been grouped together for clarity, e.g care or nursing homes.

| **Organisation** | **N reports received** | **responses (n)** | **Response rate** | **Early or on time responses** | **Late responses** | **Overdue** | **Undated (%)** |
| --- | --- | --- | --- | --- | --- | --- | --- |
| **NHS Organisations** | **249** | **144** | **58%** | **43%** | **13%** | **42%** | **2%** |
| NHS Trusts/Hospitals | 158 | 90 | 57% | 44% | 10% | 42% | 3% |
| NHS England | 24 | 12 | 50% | 21% | 29% | 50% | 0% |
| GPs | 19 | 10 | 53% | 42% | 11% | 47% | 0% |
| CCGs | 15 | 9 | 60% | 40% | 20% | 40% | 0% |
| Welsh Health Boards | 13 | 9 | 69% | 54% | 15% | 31% | 0% |
| Ambulance Services | 11 | 10 | 91% | 73% | 18% | 9% | 0% |
| Community health organisations | 6 | 2 | 33% | 33% | 0% | 67% | 0% |
| Other urgent care organisations | 2 | 1 | 50% | 0% | 0% | 50% | 50% |
| Mental Health clinics/trusts | 1 | 1 | 100% | 100% | 0% | 0% | 0% |
| **Care organisations** | **30** | **16** | **53%** | **33%** | **10%** | **47%** | **10%** |
| Care or Nursing home | 22 | 11 | 50% | 36% | 5% | 50% | 9% |
| Care providers | 8 | 5 | 63% | 25% | 25% | 38% | 13% |
| **Government bodies or depts** | **78** | **40** | **51%** | **18%** | **27%** | **49%** | **6%** |
| DHSC | 42 | 21 | 50% | 10% | 38% | 50% | 2% |
| Local government organisations | 15 | 7 | 47% | 33% | 7% | 53% | 7% |
| MHRA | 11 | 7 | 64% | 36% | 18% | 36% | 9% |
| Other national government | 6 | 4 | 67% | 17% | 17% | 33% | 33% |
| PHE | 4 | 1 | 25% | 0% | 25% | 75% | 0% |
| **Professional bodies** | **46** | **22** | **48%** | **43%** | **2%** | **52%** | **2%** |
| Royal Colleges/Societies | 22 | 13 | 59% | 50% | 5% | 41% | 5% |
| CQC | 15 | 3 | 20% | 20% | 0% | 80% | 0% |
| NICE | 4 | 3 | 75% | 75% | 0% | 25% | 0% |
| GMC | 3 | 1 | 33% | 33% | 0% | 67% | 0% |
| Other professional groups | 2 | 2 | 100% | 100% | 0% | 0% | 0% |
| **Others** | **18** | **7** | **39%** | **28%** | **11%** | **61%** | **0%** |
| Private companies | 11 | 4 | 36% | 27% | 9% | 64% | 0% |
| Universities and medical schools | 3 | 2 | 67% | 33% | 33% | 33% | 0% |
| Unaffiliated Private individuals | 1 | 0 | 0% | 0% | 0% | 100% | 0% |
| Prisons | 1 | 0 | 0% | 0% | 0% | 100% | 0% |
| Private hospitals | 1 | 1 | 100% | 100% | 0% | 0% | 0% |
| Unclear recipient | 1 | 0 | 0% | 0% | 0% | 100% | 0% |
| **Total** | **421** | **229** | **54%** | **37%** | **14%** | **45%** | **4%** |

NHS: National Health Service, GPs: general practitioners, CCGs: clinical commissioning groups, DHSC: Department for Health and Social Care, MHRA: Medicines and Healthcare Products Regulatory Agency, PHE: Public Health England, CQC: Care Quality Commission, NICE: National Institute for Health and Care Excellence, GMC: General Medical Council

**Supplementary Table S11:** Broad responses received from organisations that responded to PFDs

| **Organisation** | **N responses** | **% Responds, Acknowledges concern and initiates new change to address concern** | **(%) Pre-existing solution adequate** | **% Responds but does not acknowledge/agree with concern** |
| --- | --- | --- | --- | --- |
| **NHS Organisations** | **144** | 85% | 10% | 4% |
| CCGs | 9 | 89% | 11% | 0% |
| NHS England | 12 | 67% | 33% | 0% |
| Ambulance Services | 10 | 80% | 10% | 10% |
| NHS Trusts/Hospitals | 90 | 86% | 9% | 6% |
| GPs | 10 | 90% | 10% | 0% |
| Welsh Health Boards | 9 | 100% | 0% | 0% |
| Mental Health clinics/trusts | 1 | 100% | 0% | 0% |
| Community health organisations | 2 | 100% | 0% | 0% |
| Other urgent care organisations | 1 | 100% | 0% | 0% |
| **Care organisations** | **16** | **75%** | **19%** | **6%** |
| Care or Nursing home | 11 | 73% | 18% | 9% |
| Care providers | 5 | 80% | 20% | 0% |
| **Government bodies or depts** | **40** | **45%** | **48%** | **8%** |
| DHSC | 21 | 48% | 52% | 0% |
| Local government organisations | 7 | 71% | 29% | 0% |
| MHRA | 7 | 0% | 71% | 29% |
| Other national government | 4 | 50% | 25% | 25% |
| PHE | 1 | 100% | 0% | 0% |
| **Professional bodies** | **22** | **50%** | **41%** | **9%** |
| NICE | 3 | 33% | 67% | 0% |
| GMC | 1 | 100% | 0% | 0% |
| CQC | 3 | 0% | 100% | 0% |
| Royal Colleges/Societies | 13 | 54% | 31% | 15% |
| Other professional groups | 2 | 100% | 0% | 0% |
| **Others** | **7** | **86%** | **0%** | **14%** |
| Private companies | 4 | 75% | 0% | 25% |
| Universities and medical schools | 2 | 100% | 0% | 0% |
| Private hospitals | 1 | 100% | 0% | 0% |
| **Total** | **229** | **74%** | **20%** | **6%** |

**Supplementary Table S12:** Types of new changes implemented by organisations in response to PFDs. Percentages are expressed as a proportion of all responses received.

| **Organisation** | **N responses** | **% Improve patient info** | **% Improved training** | **% audit or investigation** | **% improve communication or handover** | **% Increase staffing** | **% New protocol, pathway, guidance document introduced** | **% Changes to record-keeping** | **% Investment or increase in resources** |
| --- | --- | --- | --- | --- | --- | --- | --- | --- | --- |
| **NHS Organisations** | **144** | **8%** | **42%** | **43%** | **19%** | **15%** | **65%** | **22%** | **13%** |
| CCGs | 9 | 0% | 44% | 56% | 0% | 0% | 78% | 11% | 22% |
| NHS England | 12 | 0% | 17% | 25% | 0% | 0% | 17% | 8% | 8% |
| Ambulance Services | 10 | 0% | 20% | 10% | 10% | 20% | 60% | 10% | 20% |
| NHS Trusts/Hospitals | 90 | 10% | 44% | 46% | 22% | 18% | 69% | 26% | 13% |
| GPs | 10 | 20% | 60% | 50% | 20% | 10% | 60% | 30% | 10% |
| Welsh Health Boards | 9 | 11% | 67% | 67% | 33% | 22% | 78% | 22% | 11% |
| Mental Health clinics/trusts | 1 | 0% | 0% | 0% | 100% | 100% | 100% | 0% | 0% |
| Community health organisations | 2 | 0% | 50% | 50% | 0% | 0% | 50% | 50% | 0% |
| Other urgent care organisations | 1 | 0% | 0% | 0% | 0% | 0% | 100% | 0% | 0% |
| **Care organisations** | **16** | **0%** | **63%** | **38%** | **19%** | **6%** | **56%** | **25%** | **6%** |
| Care or Nursing home | 11 | 0% | 73% | 36% | 18% | 9% | 45% | 36% | 9% |
| Care providers | 5 | 0% | 40% | 40% | 20% | 0% | 80% | 0% | 0% |
| **Government bodies or depts** | **40** | **3%** | **23%** | **23%** | **0%** | **3%** | **28%** | **5%** | **0%** |
| DHSC | 21 | 0% | 19% | 24% | 0% | 5% | 29% | 5% | 0% |
| Local government organisations | 7 | 0% | 29% | 43% | 0% | 0% | 43% | 14% | 0% |
| MHRA | 7 | 14% | 29% | 0% | 0% | 0% | 0% | 0% | 0% |
| Other national government | 4 | 0% | 25% | 25% | 0% | 0% | 25% | 0% | 0% |
| PHE | 1 | 0% | 0% | 0% | 0% | 0% | 100% | 0% | 0% |
| **Professional bodies** | **22** | **0%** | **5%** | **18%** | **0%** | **0%** | **36%** | **5%** | **5%** |
| NICE | 3 | 0% | 0% | 0% | 0% | 0% | 67% | 0% | 0% |
| GMC | 1 | 0% | 0% | 100% | 0% | 0% | 0% | 0% | 0% |
| CQC | 3 | 0% | 0% | 33% | 0% | 0% | 0% | 0% | 0% |
| Royal Colleges/Societies | 13 | 0% | 8% | 8% | 0% | 0% | 46% | 8% | 0% |
| Other professional groups | 2 | 0% | 0% | 50% | 0% | 0% | 0% | 0% | 50% |
| **Others** | **7** | **0%** | **57%** | **43%** | **0%** | **0%** | **57%** | **14%** | **14%** |
| Private companies | 4 | 0% | 50% | 50% | 0% | 0% | 50% | 25% | 25% |
| Universities and medical schools | 2 | 0% | 50% | 0% | 0% | 0% | 50% | 0% | 0% |
| Private hospitals | 1 | 0% | 100% | 100% | 0% | 0% | 100% | 0% | 0% |
| **Total** | **229** | **6%** | **37%** | **37%** | **13%** | **10%** | **55%** | **17%** | **10%** |

**Paediatric sub-group analyses**

**Supplementary Table S13.** ICD-11 codes for cause of death accounting for ≥0.5% cases (Section 1A of death certificate), among the paediatric sub-group.

| **ICD-11 coding for cause of death** | **Number of cases (N)** | **Percentage of cases (%)** | **Specific** |
| --- | --- | --- | --- |
| 1G40 | 14 | 45.16% | Sepsis without septic shock |
| 1G41 | 5 | 16.13% | Sepsis with Septic Shock |
| 1C1C | 3 | 9.68% | Meningococcal disease |
| MA15.0 | 3 | 9.68% | Bacteraemia |
| MG4A | 1 | 3.23% |  |
| 8B24 | 1 | 3.23% |  |
| 1E31 | 1 | 3.23% |  |
| 1C41 | 1 | 3.23% |  |
| 3B62.Y | 1 | 3.23% |  |
| KB24 | 1 | 3.23% |  |
| XN5PZ | 1 | 3.23% |  |
| XN6P4 | 1 | 3.23% |  |

**Supplementary Table S14.** Specific pathogens identified within paediatric PFDs as the causative agent in sepsis. A single PFD may mention more than one pathogen.

| **Pathogen** | **N** | **% of PFDs which named a pathogen** |
| --- | --- | --- |
| E.coli | 6 | 29% |
| N.meningitidis | 4 | 19% |
| Group A Strep | 3 | 14% |
| Group B Strep | 2 | 10% |
| Influenza | 2 | 10% |
| Pneumococcus | 2 | 10% |
| Stentotrophomonas maltophilia | 1 | 5% |
| Herpes simplex | 1 | 5% |
| **Total** | **21** | **100%** |

**Supplementary Table S15.** Source of infection as outlined in paediatric PFD reports. *Note: Some cases had more than one site of infection specified.

| **Source of Infection*** | **Number of Cases (N)** | **Percentage of cases (%)** |
| --- | --- | --- |
| Pneumonia, Other Lung Infection | 9 | 29.03% |
| UTI, Pyelonephritis | 4 | 12.90% |
| Congenital | 4 | 12.90% |
| ENT | 2 | 6.45% |
| Fracture/Injury | 1 | 3.23% |
| Abdomen (GI, Hepato-biliary) | 1 | 3.23% |
| Others | 1 | 3.23% |
| None Specified | 11 | 35.48% |

**Supplementary Table S16:** Concerns raised by coroners in line with the Royal College of Emergency Medicine (RCEM) sepsis audit for paediatric PFDs

| **RCEM category** | **Number of cases** | **% of cases** |
| --- | --- | --- |
| Issues with education and training for early recognition and instigation of optimal care | 12 | 39% |
| Issues with sepsis protocol / sepsis lead | 9 | 29% |
| Issues with senior review / escalation | 9 | 29% |
| Issues with NEWS scores and sepsis screening | 9 | 29% |
| Issues with administration of antibiotics | 6 | 19% |
| Issues with sepsis care pathways | 6 | 19% |
| Issues with timely observations | 3 | 10% |
| Issues with providing patients with written information | 3 | 10% |
| Issues with monitoring of parameters including lactate and urine output | 1 | 3% |
| Issues with administration of oxygen | 0 | 0% |

**Supplementary Table S17:** Coroners’ concerns in paediatric PFDs, grouped following thematic analysis.

| **Concern** | **Number** | **Percent of reports** |
| --- | --- | --- |
| Failure to use sepsis screening tools ie. problems with early warning systems | 8 | 26% |
| Failure to provide appropriate Treatment | 8 | 26% |
| Failure to follow relevant pathways, protocols, guidelines or risk assessments once sepsis had been recognised | 7 | 23% |
| Failure in communication or handover | 7 | 23% |
| Lack of training | 7 | 23% |
| Failure to recognise risk factors/comorbidities | 5 | 16% |
| Failure to keep accurate records / notes | 5 | 16% |
| Failure to take observations | 4 | 13% |
| Failure to escalate | 4 | 13% |
| Failure in timely medical assessment of patient | 3 | 10% |
| Poor discharge planning | 3 | 10% |
| Poor response time of emergency services | 2 | 6% |
| Understaffing | 2 | 6% |
| Lack of resources | 2 | 6% |
| Poor response time of inpatient medical staff | 1 | 3% |
| Poor response time of care home staff | 0 | 0% |
| Failure to secure imaging in timely manner/at all | 0 | 0% |
| Concern not relevant to sepsis e.g. DNAR in place was ignored | 16 | 52% |
| **Total** | **84** | **100%** |

**Supplementary Table S18.** Organisations to whom paediatric PFDs were addressed.

| **Organisation** | **N reports sent** |
| --- | --- |
| CCGs | 4 |
| NHS England | 6 |
| Ambulance Services | 2 |
| NHS Trusts/Hospitals | 13 |
| GPs | 1 |
| Welsh Health Boards | 2 |
| Other urgent care organisations | 1 |
| DHSC | 8 |
| MHRA | 1 |
| Other national government | 2 |
| NICE | 4 |
| CQC | 4 |
| Royal Colleges/Societies | 4 |
| Other professional groups | 1 |
| **Total** | **53** |

**Supplementary Table S19.** Response rate of organisations in receipt of a paediatric sepsis-related PFD.

| **N reports received** | **responses to reports (n)** | **Response rate** | **Early or on time responses (%)** | **Late responses (%)** | **Overdue %** | **Undated (%)** |
| --- | --- | --- | --- | --- | --- | --- |
| 53 | 21 | 40% | 23% | 15% | 60% | 2% |

**Supplementary Table S20.** Broad response theme of organisations in receipt of a paediatric sepsis-related PFD

| **N responses** | **% Responds, acknowledges concern and initiates new change to address concern** | **(%) Pre-existing solution adequate** | **% Responds but does not acknowledge/agree with concern** |
| --- | --- | --- | --- |
| 21 | 62% | 33% | 5% |

**Supplementary Table S21.** Specific responses by organisations initiating a new change in response to a paediatric sepsis-related PFD

| **N responses** | **21** |
| --- | --- |
| **% Improve patient info** | 5% |
| **% Improved training** | 29% |
| **% audit or investigation** | 38% |
| **% improve communication or handover** | 10% |
| **% Increase staffing** | 10% |
| **% New protocol, pathway, guidance document introduced** | 52% |
| **% Changes to record-keeping** | 24% |
| **% Investment or increase in resources** | 10% |
